# Exploring and accounting for genetically driven effect heterogeneity in Mendelian Randomization

**DOI:** 10.1101/2024.05.05.24306667

**Authors:** Annika Jaitner, Krasimira Tsaneva-Atanasova, Rachel M. Freathy, Jack Bowden

## Abstract

Mendelian randomization uses genetic variants as instrumental variables to estimate the causal effect of a modifiable health exposure or drug target on a downstream outcome. A crucial assumption for accurate estimation of the average causal effect in MR, termed Homogeneity, is that the causal effect does not vary across levels of any instrument used in the analysis. In contrast, the science of pharmacogenetics seeks to actively uncover and exploit genetically driven effect heterogeneity for precision medicine. In this paper we consider a recently proposed method for performing pharmacogenetic analysis on observational data (The Triangulation WIthin a STudy (TWIST) framework), and explore how it can be combined with traditional MR approaches to properly characterise average and causal effects and genetically driven effect heterogeneity. We propose two new methods which not only estimate the genetically driven effect heterogeneity but also enable the estimation of a causal effect in the genetic group with and without the risk allele separately. Both methods utilise homogeneity-respecting and homogeneity-violating genetic variants and rely on a different set of assumptions. Using data from the ALSPAC study, we apply our new method to estimate the causal effect of smoking before and during pregnancy on offspring birth weight in mothers whose genetics mean they find it (relatively) easier or harder to quit.

## 1 Introduction

Confirming or refuting causal relationships is difficult in observational study settings as one can never be sure if all confounders have been identified, appropriately measured and adjusted for. However, one can take advantage of random genetic inheritance from parents to offspring in an observational analysis to help uncover true causal mechanisms and estimate the causal effect of health interventions [1]. Mendelian randomization (MR) is the formal science of using genetic variants as instrumental variables (IV) for this purpose [2]. Rather than testing the direct association between an exposure and outcome, a genetically predicted exposure is used instead. Under the assumption of random distribution of genetic variants from parents to offspring at conception, an individual’s genetically predicted exposure should be far less susceptible to confounding bias. MR requires three core assumptions to hold for a genetic variant, *G*, to be valid instrument to test for a causal relationship between a modifiable exposure and health outcome [3]. These are termed the relevance assumption, the independence assumption and the exclusion restriction. To go beyond testing for causality, an additional assumption is required to estimate (or ‘point identify’) the causal effect. The most commonly used fourth assumption is termed Homogeneity. It states that the causal effect an individual experiences is not affected by the value of their genetic instrument. When this is satisfied, an IV analysis can in theory estimate the average causal effect of intervention on the exposure for an entire population. However, for continuous outcomes this assumption is often biologically implausible unless a suitable ‘typical’ range for the exposure is defined [4]. In cases where the Homogeneity assumption is deemed implausible, an alternative assumption termed Monotonicity can instead be applied to enable causal estimation [5]. In the context of an MR study using a genetic variant *G*, Monotonicity means that there is no individual whose exposure would be higher if they did not carry the exposure raising allele of *G* than if they did. Such individuals are termed ‘Defiers’, and assuming that none exist allows the estimation of the causal effect in the genetic ‘Complier’ subset - defined as the group of individuals whose exposure level would always be greater with the exposure-raising allele of *G* than without.

Although Homogeneity is typically a baseline assumption for most MR studies, in pharmacogenetic investigations genetic variants are explicitly sought to explain apparent heterogeneity in a treatment’s effectiveness. For example, many pharmaceutical interventions are pro drugs, which require a specific metabolic process to occur for the patient to experience the full treatment effect. I f t he p atient h as a g enetic v ariant that hinders the drug’s metabolism (e.g. a ‘Loss-of-function’ (LoF) mutation), the treatment effect m ay b e less pronounced in individuals who carry it. For example, Pilling et al. [6] showed that *CYP2C19* LoF alleles were associated with higher incidence of ischaemic events amongst those taking the commonly prescribed anti-stroke drug, Clopidogrel. National Institute of Health and Care Excellence guidance now recommends genotyping individuals on Clopidogrel who experience an ischaemic event to be genotyped, with a view to altering their medication if the LoF variant is found [7].

Observational data can be used to quantify the extent of genetically driven treatment effect heterogeneity, but the analysis can be compromised by strong confounding by indication and off-target genetic effects on the outcome of interest that are independent of any gene-drug interaction. A recently proposed method of pharmacogenetic causal inference using observational data - Triangulation Within a Study (TWIST)[5] - defined the assumptions required to estimate the difference in treatment effect estimates between those with and without a pharmacogenetic variant, as a measure of genetically driven effect h eterogeneity. A range of different m ethods were proposed to e stimate this q uantity as well as a f ramework f or c ombining them if sufficiently si milar. Although it is a useful tool for estimating this difference, in its most basic form it cannot estimate the causal effect of treatment on the outcome in each genetic group, which is a limitation.

Instances of genetically driven effect heterogeneity do exist in main stream epidemiological investigations of non-pharmaceutical interventions. For example, smoking in pregnancy has been shown to have measurable consequences on offspring b irth w eight, w hich i s a n i mportant m arker o f l ong-term h ealth [8]. Specifically, Freathy et al. [9] show that single-nucleotide polymorphism (SNP) rs1051730 on chromosome 15 is associated with smoking cessation during pregnancy as well as smoking quantity. However, the same SNP is not associated with smoking initiation. Therefore, mothers with the rs1051730 risk allele are not more likely to smoke than mothers without, but if they do smoke they tend to smoke more heavily than non-carriers and find it harder to quit, meaning the effect of smoking on birth weight could easily be moderated by rs1051730.

In this paper we review the standard MR method, which utilises homogeneity-respecting genetic instruments, and the TWIST method, which utilises homogeneity-violating instruments. We highlight the different conceptual starting points for each approach, in terms of their modelling assumptions, and how estimates are biased if these assumptions are violated. Subsequently, we explore the integration of both sets of instruments into a unified analysis in order to properly characterise the average causal effects and genetically driven effect heterogeneity. Using data from the ALSPAC study, we apply our new method to estimate the causal effect of smoking on offspring birth weight in distinct genetic subgroups of pregnant mothers; the magnitude of the effect heterogeneity; and the potential public health impact of genetically targeted treatment going forward.

## 2 Methods

Let *S* and *G* be binary variables capturing the exposure and genetic variant of interest. In our applied example *S* reflects the smoking status of the m other. We allow for the effect of the exposure on the outcome, Y, to be altered through an interaction with *G*, denoted as *S*^*\**^ = *G* × *S*. To motivate the method, we assume the following linear interaction model for the mean outcome *Y* given *S, G*, and additionally measured (*Z*) and unmeasured (*U*) confounders of *S* and *Y* respectively:

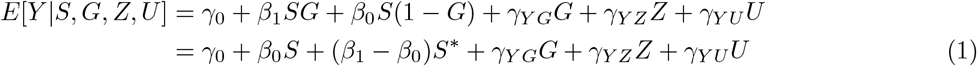

Figure 1 (a) depicts the directed acyclic graph consistent with the model described in equation (1) and highlights various key assumptions using coloured arrows. We first consider the traditional set of assumptions required in order to estimate the average causal effect (ACE) of the exposure on the outcome. We can express the ACE as the expected contrast between the potential outcomes of all mothers if they smoked during pregnancy and if they did not

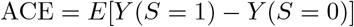

**Figure 1:**
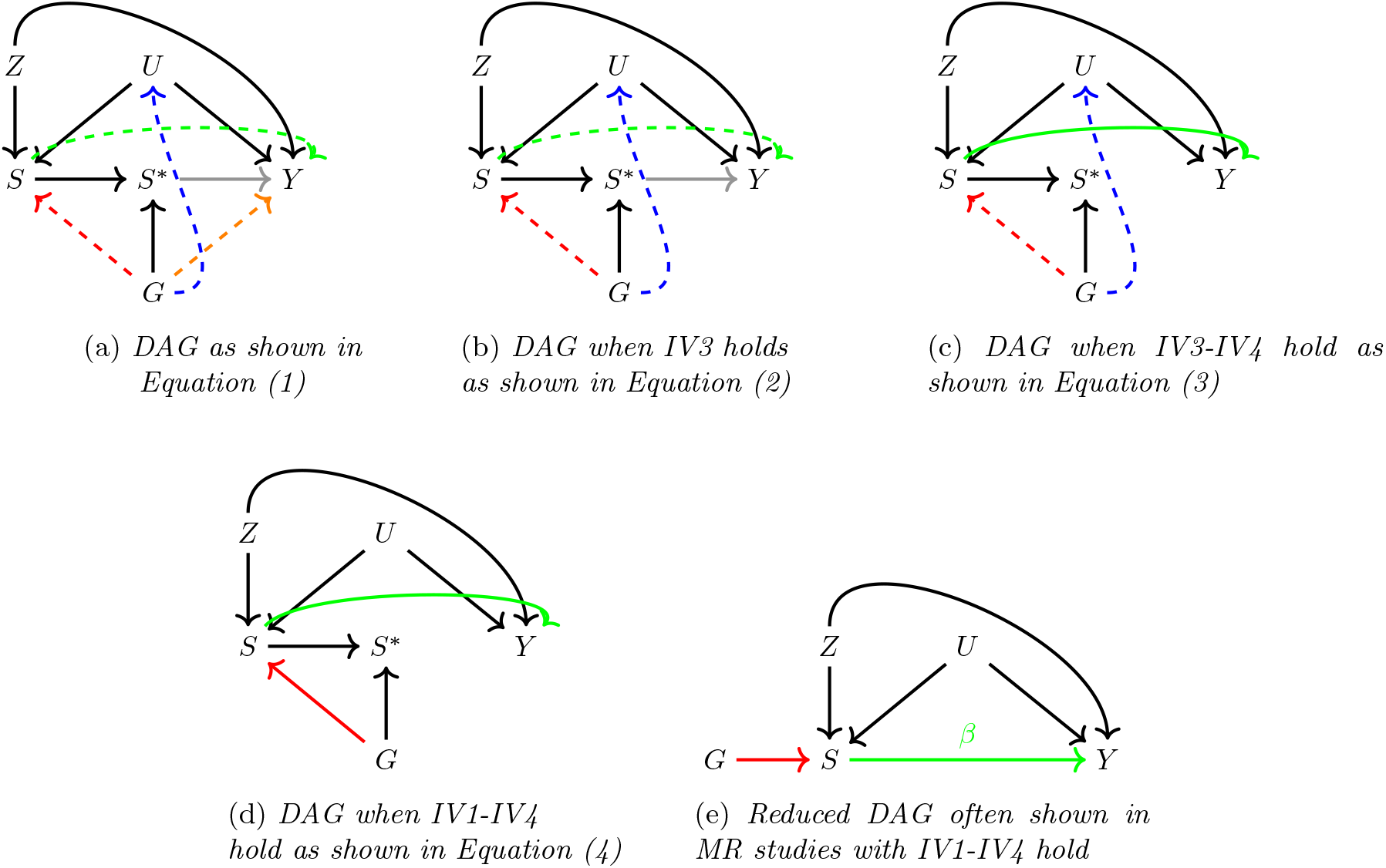
Diagram reduces step wise with respect to IV1-IV4 assumptions.

These assumptions are [10]

IV1 (relevance) : The genetic instrument *G* predicts the exposure *S* (a red arrow);

IV2 (independence) : The genetic instrument *G* is independent of any confounders *U* (**no** blue arrow);

IV3 (exclusion) : The genetic instrument *G* is independent of the outcome *Y* given the exposure *S* and any confounders *U* (**no** orange arrow).

IV4 (homogeneity) : The effect of the exposure *S* on the outcome *Y* is independent of the genetic instrument *G* (**no** grey arrow).

Assumptions IV1-4 enable us to extract the ACE via an IV analysis, by turning the general model and causal diagram in Figure 1 (a) into the reduced model and causal diagram in Figure 1 (e), through the following steps:

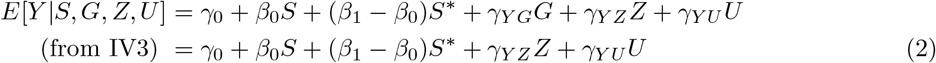

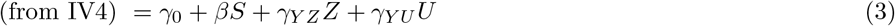

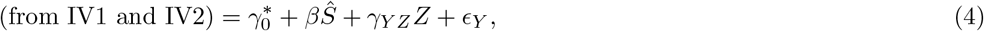

where *Ŝ* = *E*[*S*|*G*] and *ϵ*_*Y*_ is a residual error term that is crucially independent of *Ŝ*. The reduced causal diagram in Figure 1 (e) is often shown in MR studies.

### 2.1 What does an MR analysis estimate under violation of IV2-4?

We now consider what the MR estimator targets, assuming the data model described in equation (1), when IV1 holds but initially, assumptions IV2-IV4 do not. Under our assumed model as described in equation (1), the MR estimator is as follows:

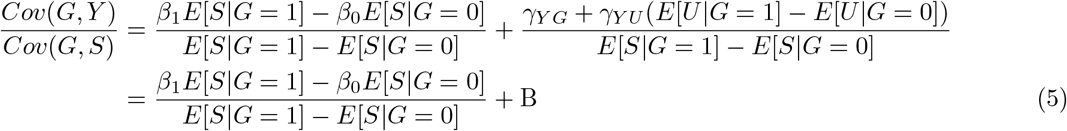

IV1 guarantees that the denominator of (5) is non-zero and so the ratio terms are well defined. The second term in (5), *B*, represents the bias due to violation of IV2 and IV3. If Homogeneity, but Monotonicity holds, we show in *Supplementary material* that equation 5 equals the Complier Average Causal Effect (CACE) plus any bias due to violation of IV2 and IV3. Compliers are defined as individuals that smoke i f they have the risk allele (*G* = 1) and do not smoke if they do not have it (*G* = 0).

### 2.2 Genetically moderated exposure effect (GMEE)

Genetic instruments that satisfy the Homogeneity assumption enable estimation of the ACE. However, in studies into the consequences of smoking versus not smoking, this assumption will be demonstrably false if attempting the analysis with a SNP like rs1051730, since the smoking patterns of people with and without this variant are likely to be different. I n this c ase, a m ore practical s tarting p oint would b e to assume the underlying DAG structure in Figure 1 (left) and aim to quantify the magnitude of Homogeneity violation as the difference in smoking effects between the two genetic su b-groups. This ‘genetically moderated exposure effect’ (GMEE) i s represented by arrow b etween *S* ^*\**^ and the outcome *Y*. From equation (1) this i s equal to *β*_1_ − *β*_0_.

Bowden et al. [5] discuss various methods for estimating this quantity, which we refer to as the GMEE, but which they referred to as the GMTE (T being for treatment). Each of the methods presented in Bowden et al. [5] relies on a different s et o f a ssumptions. For e xample, w hen t he g enetic i nstrument *G* i s independent of the exposure (i.e. no red arrow in Figure 1 due to violation of IV1), independent any unmeasured confounder (i.e. no blue arrow in Figure 1, IV2 satisfied), and only affects the outcome through the moderated exposure variable (i.e. no orange arrow in Figure 1, IV3 satisfied), the GMEE can be estimated in the exposed population only. In our setting, this would be estimated by the difference in mean outcomes across the genetic groups amongst the population of smokers only:

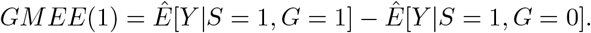

Here the ‘(1)’ notation reminds the analyst that only smoker’s data is used and *G* = 1*/*0 refers to the presence/absence of at least one risk allele of SNP rs1051730. A more robust estimate of the GMEE is the

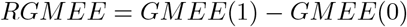

where

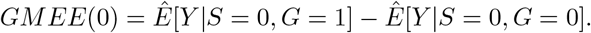

Here, the R prefix in RGMEE stands for ‘robust’, since i t can estimate the GMEE without bias even i f IV3 is violated (*G* affects the o utcome d irectly a s i ndicated by the o range a rrow i n Figure 1). I ndeed i t i s this bias term that is estimated by GMEE(0) before being subtracted out.

Bowden et al. [5] state that the RGMEE is unbiased even if the genetic instrument violates IV2, by being associated with the outcome through the unmeasured confounder (blue arrow in Figure 1. Our investigations in this paper have shown this to be incorrect (See our erratum [11]). Nevertheless, this means we are now able to straightforwardly verify if the assumptions for the RGEE (i.e. a desired violation of IV1 but no violation of IV2) hold, since they imply that *G* and *S* are independent. Testing for an association between *G* and *S* is therefore an important pre-requisite for its use.

## 3 Enhancing robustness through the integration of MR and GMEE methods

The genetically moderated exposure effect introduced in the previous section proposes an array of methods for estimating the difference *β*_1_ − *β*_0_ under different assumptions, but not the individual values *β*_1_ and *β*_0_. To address this, we now formally extend the previous framework by incorporating a second variant, *G*_2_, that is a ‘standard’ instrument for the exposure satisfying assumptions IV1-4. In our case, it therefore influences smoking initiation directly, but does not moderate an individual’s smoking habits, thereby violating Homogeneity. We now explore two scenarios that expand upon the standard TWIST approach, utilising novel methods that leverage the two available genetic instruments. The DAGs for these two separate methods are shown in Figure 2.

**Figure 2:**
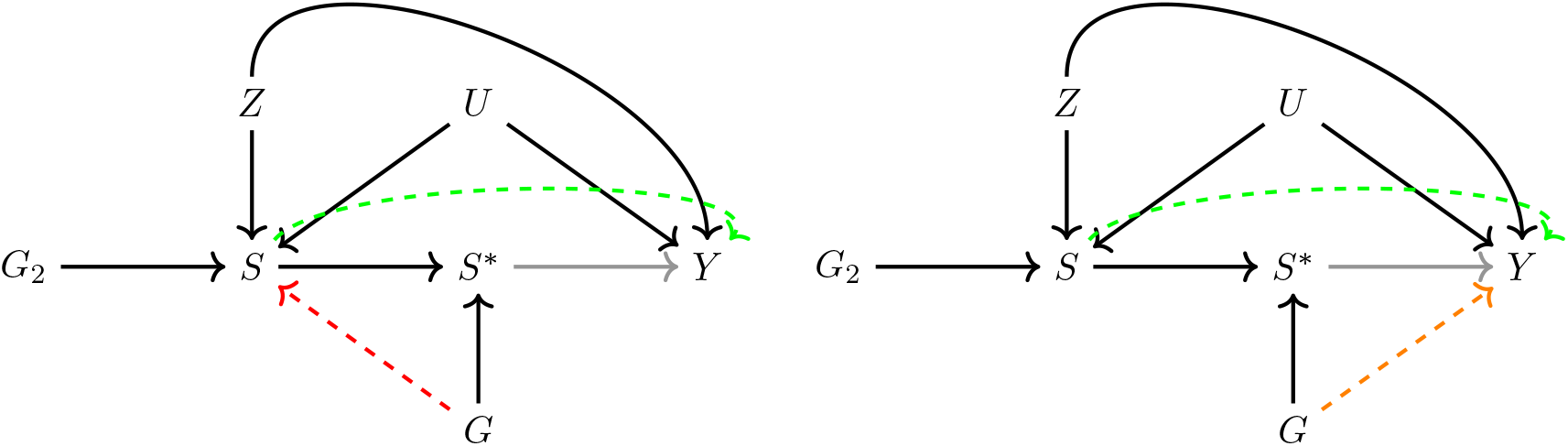
Left: DAG of linear interaction model from equation (1) highlighting in colour which assumptions need to hold for an unbiased estimate of β_1_ and β_0_ when using Method 1. Right: DAG of linear interaction model (1) highlighting in colour which assumptions need to hold for an unbiased estimate of β_1_ and β_0_ when using Method 2.

### 3.1 Method 1: (*G, G*_2_) are jointly valid instruments for (*S,S*^*\**^)

We first consider estimation of *β*_1_ and *β*_0_ using both genetic instruments *G* and *G*_2_ within a multivariable model. This can be enacted in a two-step procedure by using *G* and *G*_2_ to predict *S* in stage 1, and then plugging in the predicted values into the linear interaction model in stage 2:

1. Stage 1: Estimate *Ŝ* = *Ê*[*S*|*G, G*_2_] using either linear or logistic regression;
2. Stage 2: *Y* = *γ*_0_ + *β*_1_*ŜG* + *β*_0_*Ŝ*(1 − *G*), with *Ŝ*: fitted values from stage 1

This approach is robust to the case where the (assumed) effect modifying variant *G* violates IV1 but satisfies IV2 and IV3 (Figure 2, Left). In *Supplementary material* we show through simulation that the true values of *β*_1_ and *β*_0_ can be recovered when these assumptions are satisfied, but violation of the assumptions lead to bias. The standard error for *β*_1_ and *β*_0_ can be obtained directly from the linear model output. As both parameters are estimated in the same model we can use the covariance matrix of *β*_1_ and *β*_0_ to derive the variance of *β*_1_ − *β*_0_ and hence can estimate the standard error for *β*_1_ − *β*_0_.

### 3.2 Method 2: Allowing for a pleiotropic effect of *G* on *Y*

We now propose a robust procedure that combines the general MR approach with the RGMEE given in Bowden et al. [5]. We first apply the RGMEE method to consistently estimate the genetically moderated effect *β*_1_ − *β*_0_. We then define a new variable *Y* (*S*^*\**^ = 0) created by subtracting the genetically moderated effect times the moderated exposure from the original outcome *Y*. More formally, *Y* (*S*^*\**^ = 0) is a potential outcome in which the treatment effect of *S*^*\**^ on *Y* has been removed. It is equal to *Y* (and therefore observed) for individuals with an *S*^*\**^=0, but is unobserved for those with *S*^*\**^=1. Finally, we perform an MR analysis using the genetic instrument *G*_2_, the exposure *S* and *Y* (*S*^*\**^ = 0). This enables estimation of *β*_0_, which can then be used in combination with the RMGEE to estimate *β*_1_.

1. Estimate the RGMEE 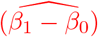 using *G*;
2. Estimate *Ŝ* = *Ê*[*S*|*G*_2_] using either linear or logistic regression;
3. Estimate *β*_0_ from model *E*[*Y* (*S*^*\**^ = 0)|*Ŝ*] = *γ*_0_ + *β*_0_*Ŝ*, where 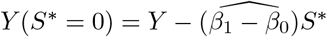

Method 2 delivers unbiased estimates if the RGMEE estimate can be consistently estimated using *G* and *β*_0_ can be consistently determined using *G*_2_ once the GMEE effect has been removed. Compared to Method 1, it allows a direct pleioptropic effect of *G* on *Y* (IV3 violation) but requires *G* to be independent of *S* (IV1 violated, but IV2 satisfied). When these assumptions are not met, our simulations show that it leads to bias (see *Supplementary Material*). The standard error for *β*_0_ and *β*_1_ − *β*_0_ can be directly taken from the respective model output. We make the assumption that 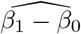 is independent of 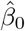, so that 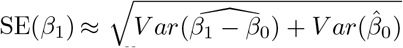. In simulations we show that it leads only to confidence intervals (CI) with a small over-coverage.

### 3.3 What does the standard MR estimate using *G*_2_ as the IV target?

When including a homogeneity respecting instrument as shown in Figure 2 a standard MR analysis with *G*_2_ as the IV is possible. Using the two-stage regression approach means:

1. Stage 1: Estimate *Ŝ* = *Ê*[*S*|*G*_2_] using a linear or logistic regression;
2. Stage 2: *Y* = *α*_0_ + *α*_1_*Ŝ* + *α*_2_*Z* + *ϵ*_*Y*_.

Here, *α*_1_ is the average causal effect on the outcome *Y* if all mothers where exposed to if all mothers were not exposed: *E*[*Y* (*S* = 1)] − *E*[*Y* (*S* = 0)]. It can be shown that under the model described in Figure 2 and equation (1):

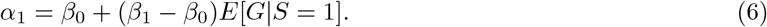

## 4 Simulation results

### 4.1 Data generation

We simulated data consistent with Figure 2 in the following manner:

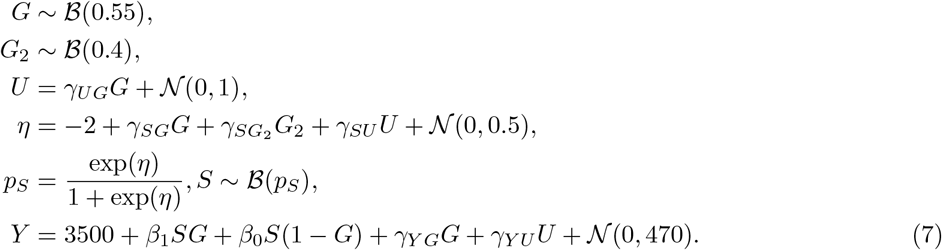

The outcome model 7 was chosen so that simulated data closely matched real birth weight data (in grams) for mothers with a history of smoking in the Avon Longitudinal Study of Parents and Children (ALSPAC) [12, 13, 14] which we will subsequently use in our applied analysis. By choosing zero and non-zero values for the parameters *γ*_*UG*_, *γ*_*SG*_ and *γ*_*Y G*_, we were able to explore the performance of Method 1 and Method 2 in estimating the causal effect parameters *β*_1_ and *β*_0_. We choose to set *β*_1_ = −200 and *β*_0_ = −100, which assumes a genetically moderated effect of *β*_1_ − *β*_0_ = − 100 grams. For all simulations we made sure that the assumptions of method 1 and 2 held. Each simulation was repeated 20,000 times (*N* = 20, 000), which enabled the calculation of bias, coverage and statistical power. For further details, Table 3 *Supplementary Material* provides a summary of the simulated data under all of the explored scenarios.

### 4.2 Estimation accuracy with increasing sample size

We investigated the sample size needed to unbiasedly estimate *β*_1_ and *β*_0_ using each approach when their respective assumptions were satisfied. Data sets were generated with sample sizes between 100 and 80,000 individuals. Figure 3 shows the mean values of *β*_1_, *β*_0_ and *β*_1_ − *β*_0_ using method 1 and method 2 across 20,000 simulations. Shaded areas reflect, for each mean parameter estimate obtained from a given sample size, a 95% confidence interval which was calculated as ±1.96 × *SD*(.) around the mean, *SD*(.) being the standard deviation of the 20,000 estimates [15]. Note that for each estimate we display three sub-figures (per column) with a different range on the y-axis: one for small sample sizes, one for medium size sample sizes and one for large sample sizes. Details on the parameter values used for each simulation are described in *Supplementary Material*. Estimation of *β*_1_ and *β*_0_ become more precise with shrinking confidence intervals as the sample size increases. Both, method 1 and 2 lead to similar results. However, for small sample sizes (below 2000), confidence intervals for method 1 estimates are wider. The third column of Figure 3 shows the mean estimates for *β*_1_ − *β*_0_ using method 1 and method 2. Here we can see a distinction in the performance of the methods across all sample sizes. Method 2, which uses the RGMEE method, yields narrower confidence intervals for 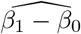 even for small sample sizes.

**Figure 3:**
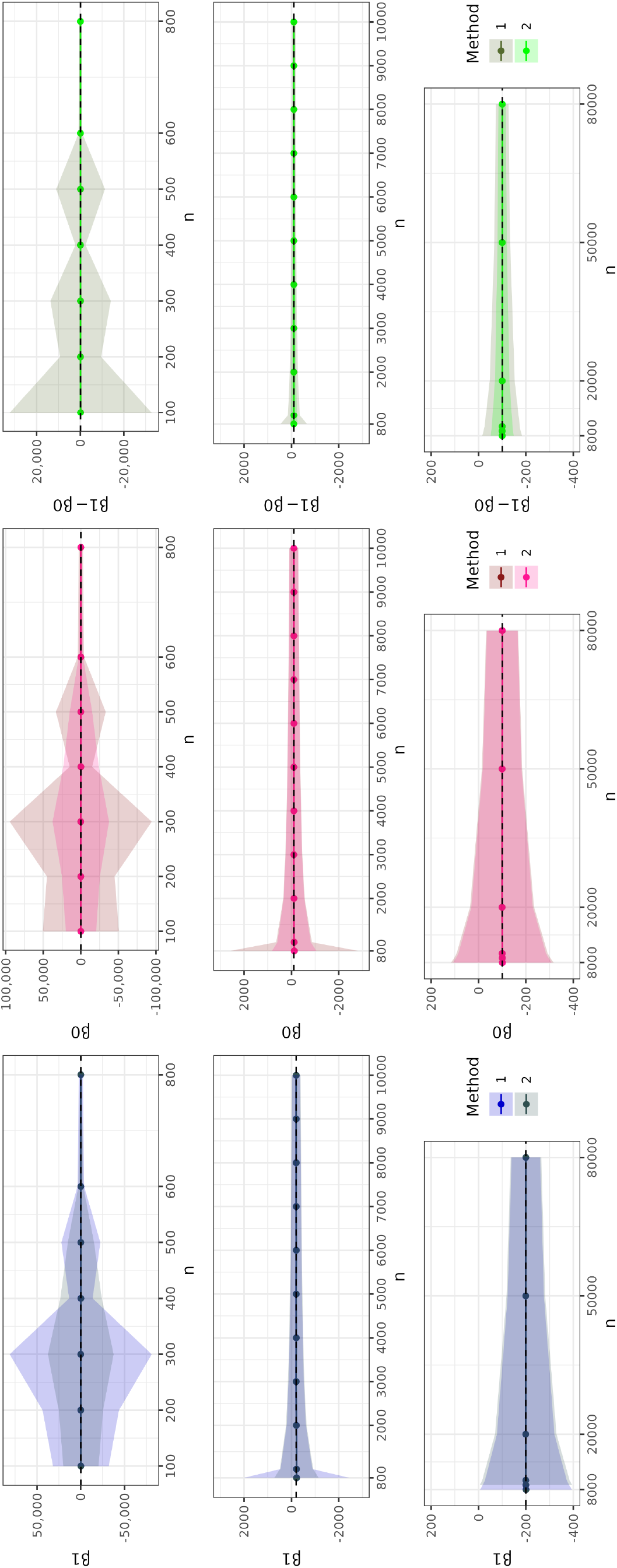
**Mean estimates** for *β*_1_, *β*_0_ and *β*_1_ − *β*_0_ over 20,000 simulations for different sample sizes. The shaded area shows the 95% CI interval derived with the Monte Carlo standard error. The dashed line indicates the true value.

### 4.3 Power and the coverage

We estimated the power to reject the null hypothesis that *β*_1_, *β*_0_ and *β*_1_ − *β*_0_ were statistically different from zero at the 5% significance level, using method 1 and 2. For each simulation we also calculated confidence intervals for the parameter estimates based on estimated standard errors, and report the coverage of 95% confidence intervals across the 20,000 simulations. The results for power and coverage are shown in Figure 4 and 5 respectively, along with their Monte-Carlo standard errors [15]. Our results show that method 2 results in a higher power when estimating *β*_0_ and *β*_1_ − *β*_0_ than method 1 for a given sample size. However, when considering estimation of *β*_1_, this is reversed. The power to detect *β*_0_ is lower than the power to detect *β*_1_ due to its lower effect size of -100 (compared to *β*_1_ = −200). Figure 5 reveals a near nominal coverage for both methods close to 95%. Crucially, our assumption that 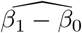 and 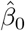 are independent leads only to a small over-coverage when estimating a confidence intervals for *β*_1_ with method 2.

**Figure 4:**
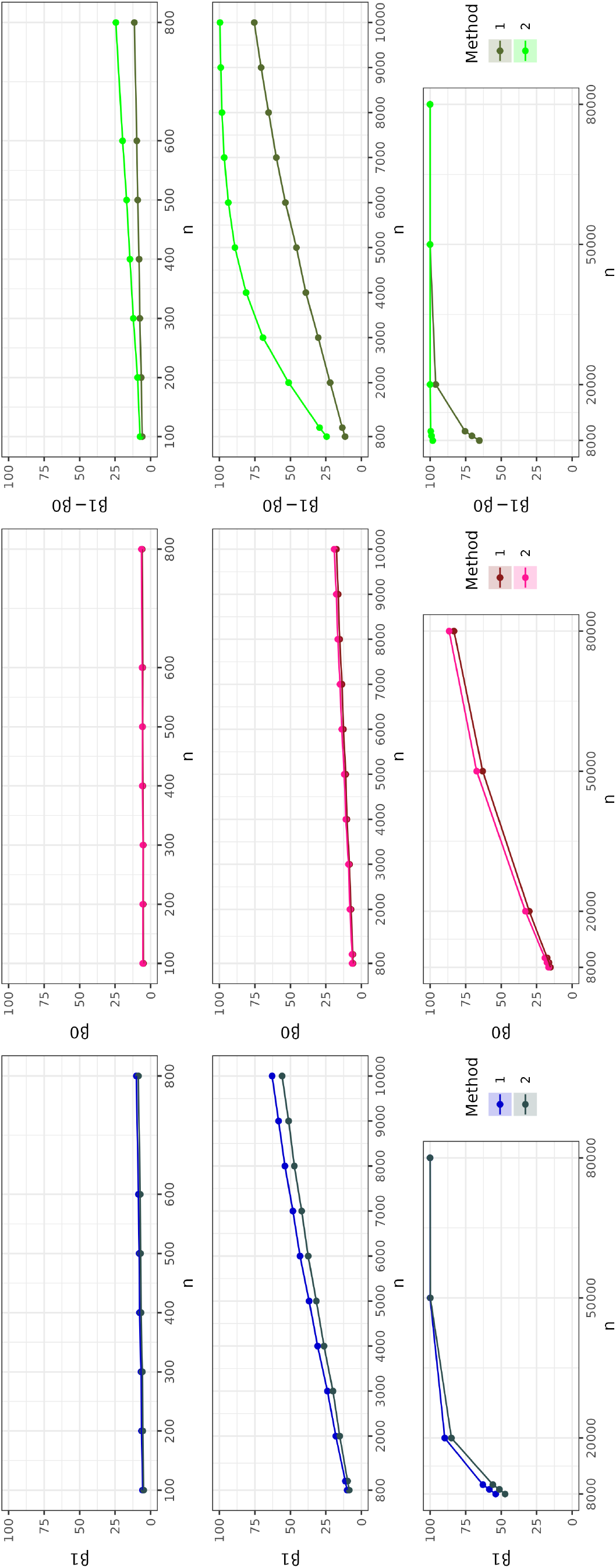
**Power** for *β*_1_, *β*_0_ and *β*_1_ − *β*_0_ over 20,000 simulations for different sample sizes. The shaded area shows the 95% CI interval for the power derived with the Monte Carlo standard error.

**Figure 5:**
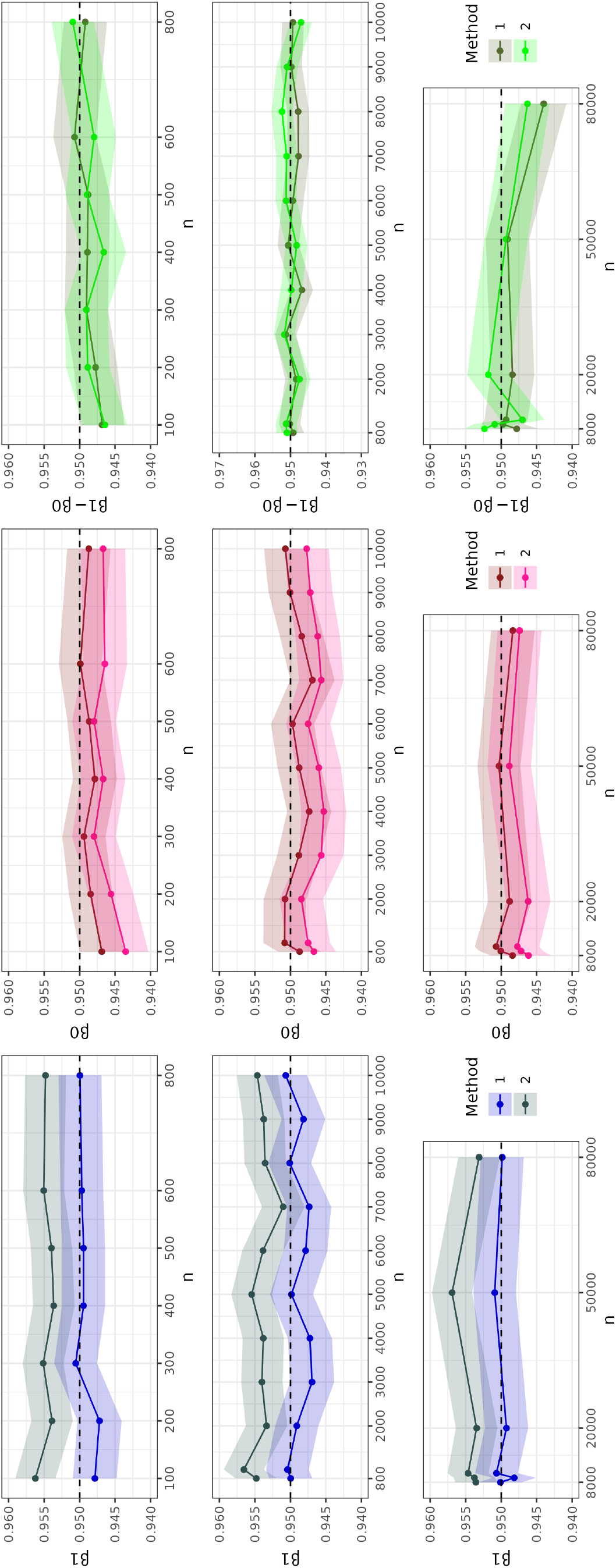
**Coverage** for *β*_1_, *β*_0_ and *β*_1_ − *β*_0_ over 20,000 simulations for different sample sizes. The shaded area shows the 95% CI interval for the coverage derived with the Monte Carlo standard error.

## 5 Applied example

### 5.1 Biological example for genetically driven exposure effects

Research into the adverse consequences of smoking has been ongoing since the 1950’s, up until the present day [16, 17]. In the specific context of maternal health, it is well established that smoking during pregnancy is associated with lower offspring birth weight, which is itself an important predictor of infant mortality and many later life health outcomes, such as cardiovascular disease, high blood pressure, coronary heart disease and type 2 diabetes [18, 19, 20]. Attributing the correct proportion of these estimated associations that are due to the causal consequences of smoking is not straightforward, due to strong confounding between smoking and later life outcomes by socio-economic factors which are very hard to completely control for. Despite this, smoking is viewed as a key modifiable risk factor, and reducing the its prevalence during pregnancy remains an important public health target [21, 17]. Unfortunately, NHS digital service statistics indicate that approximately 8.6 % of UK mothers were known smokers at the time of delivery in the first half of 2023 [22]. Identifying which individuals are at a higher risk of not giving up smoking and therefore might face more severe pregnancy outcomes can be crucial when targeting smoking cessation programmes, in order to provide support as well as closer monitoring during pregnancy.

Recently, genome-wide association studies (GWAS) have identified genetic variants that are associated with smoking initiation, smoking cessation, the age of starting smoking and smoking quantity [23]. Freathy et al. [9] show that rs1051730 on chromosome 15 is associated with smoking cessation during pregnancy as well as smoking quantity. A strong biological rationale for this exists as rs1051730 is in the nicotine acetylcholine receptor gene cluster *CHRNA5-CHRNA3-CHRNB4*. Rare variant burden associations have implicated all three of these genes as important in influencing smoking quantity [24]. However, it has also been shown that rs1051730 is *not* associated with smoking initiation [9]. The methods we have introduced thus far appear well suited to estimating the causal effect of smoking on birth weight using traditional genetic instruments for smoking initiation, whilst at the same time, quantifying the genetically moderated smoking effect via rs1051730.

### 5.2 The effect of smoking on birth weight in the ALSPAC study

The Avon Longitudinal Study of Parents and Children (ALSPAC)[12, 13, 14] invited pregnant women resident in Avon, UK with expected dates of delivery between 1st April 1991 and 31st December 1992, to take part in the study. 20,248 pregnancies have been identified as being eligible and the initial number of pregnancies enrolled was 14,541. Of the initial pregnancies, there was a total of 14,676 foetuses, resulting in 14,062 live births and 13,988 children who were alive at 1 year. We restricted our analysis to unrelated mothers with available genetic information. Additionally, we excluded multiple births and preterm births (pregnancy duration ≤ 37 weeks) [25]. The analysis data set had a sample size of 7752 individual mothers. For the traditional genetic instrument ‘*G*_2_’ we created a weighted genetic risk score (GRS) amongst the smoking initiation SNPs identified by the latest GWAS [23]. The effect sizes from the same GWAS were used as weights. We used rs1051730 as genetic effect-modifying instrument ‘*G*’, coded as 0 and 1 corresponding to no and at least one risk allele respectively. Various different smoking definitions were used for the exposure outlined in the following sections. The ALSPAC study website contains details of all the data that is available through a fully searchable data dictionary and variable search tool (http://www.bristol.ac.uk/alspac/researchers/our-data/).

#### 5.2.1 Exposure *S*= Smoking before pregnancy

Each mother was asked at 16-18 weeks of gestation whether she smoked before pregnancy. We coded mothers that reported ‘yes’ as *S* = 1 and mothers who reported ‘no’ as *S* = 0. Figure 6 (i) displays the assumed DAG for our analysis. We aimed to apply method 1 and 2 to estimate the causal effect of pre-pregnancy smoking on birth weight in the *G* = 1 group, *β*_1_, the *G*=0 group, *β*_0_, and also the genetically moderated exposure effect *β*_1_ − *β*_0_. We would expect this latter quantity to be non-zero if the pre-pregnancy smoking effect persisted differently throughout pregnancy across the two genetic groups. For the first stage of method 1, we perform a logistic regression of *S* on the GRS of smoking initiation (*G*_2_) and rs1051730 (*G*). The results are shown in Table 1. Variant rs1051730 was not associated with smoking before pregnancy, which helpfully means that method 2 is not ruled out as an analysis option. The GRS is also associated with smoking before pregnancy and we assume it acts as a true IV for this exposure. Two crucial assumptions are that the GRS of smoking initiation has no pleiotropic effect on birth weight and it does not modify the the causal effect between smoking and birth weight in the exposed and the unexposed. In order to apply method 1, rs1051730 cannot have a pleiotropic effect on birth weight either but, for method 2, this assumption is relaxed. The results from applying both methods are shown in Figure 7. To increase the precision of the estimates we adjust our regression models for different sets of covariates. The model for the genetic prediction of smoking is adjusted for whether the partner of the mother smoked, the mothers age and the first 10 genetic principal components. The model predicting birth weight is adjusted for offspring sex, mothers age, mothers height, parity, mothers pre-pregnancy weight and the first 10 genetic principal components. We viewed these variables are confounders for either smoking before pregnancy or birth weight or both. For mothers who have at least one *G* risk allele, the average causal effect of smoking before pregnancy, *β*_1_, was estimated to be a 168 g and 169 g reduction in birth weight using method 1 and method 2 respectively. On the other hand, the corresponding causal effect (*β*_0_) in mothers without a rs1051730 risk allele is a 159 g and 161 g birth weight reduction for method 1 and 2 respectively compared to non-smoking mothers without the risk allele.

**Table 1:** Logistic regression model results for logit(Pr(S = 1)) = α_0_ + α_1_G + α_2_G_2_ + δ^T^ **Z** as the first stage of method 1 using pre-pregnancy smoking as the outcome variable S. G_2_= smoking initiation GRS, G = rs1051730 and **Z** is a vector of covariates.

| S = |  | Est. | Std. Error | P-value | Fstat |
| --- | --- | --- | --- | --- | --- |
| Smoking before pregnancy | rs1051730 ($G$) | $\hat{\alpha}_1=0.02$ | 0.06 | 0.73 | 0.12 |
| | GRS smoking initiation ($G_2$) | $\hat{\alpha}_2=1.46$ | 0.16 | 4.85e-20 | 84 |
| Smoking in the first three months | rs1051730 ($G$) | $\hat{\alpha}_1=0.13$ | 0.07 | 0.06 | 3.55 |
| | GRS smoking initiation ($G_2$) | $\hat{\alpha}_2=1.39$ | 0.18 | 2.58e-15 | 62.6 |

**Figure 6:**
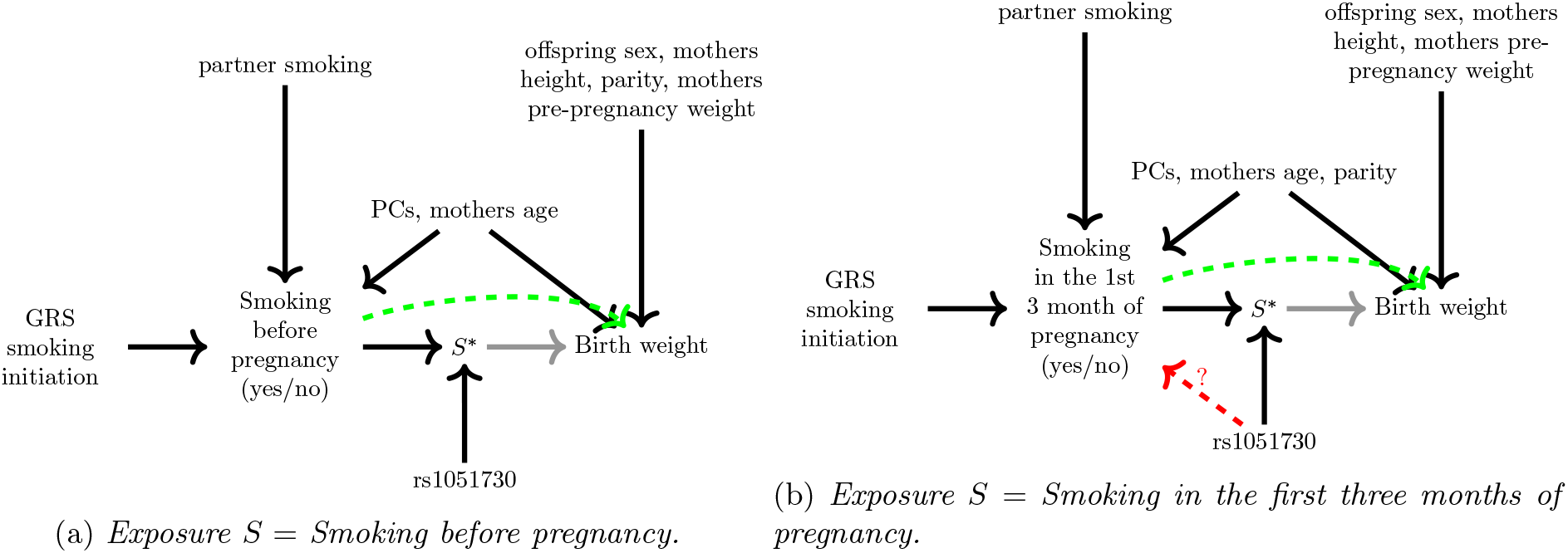
DAG showing the relationships between the genetic instrument, the exposure, the outcome and how those are affected by different confounders.

**Figure 7:**
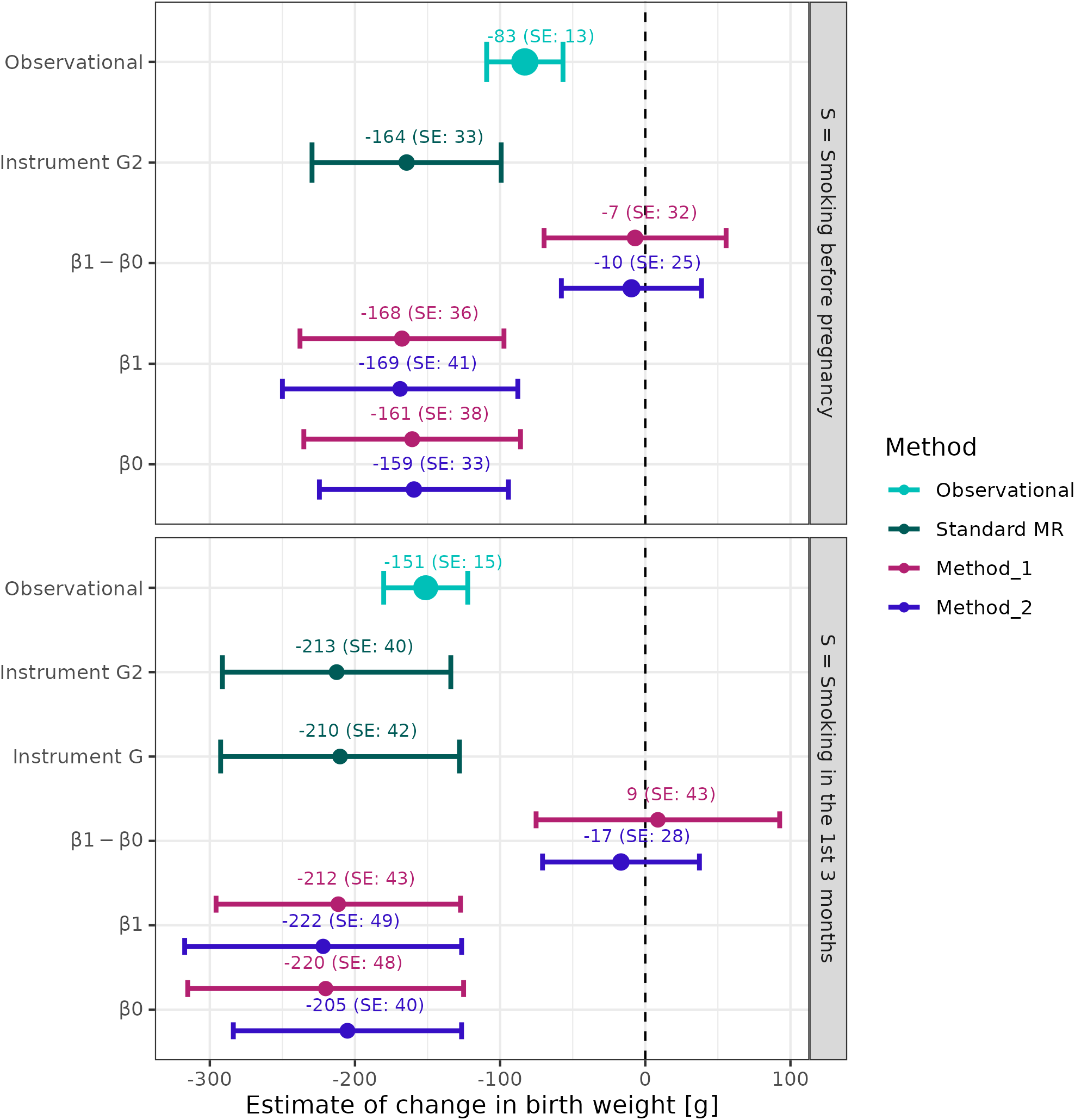
β_1_ − β_0_, β_1_ and β_0_ estimates from applying method 1 and method 2 to the ALSPAC data set. Bars indicate the 95% confidence interval. Observational analysis results arise from a linear regression of the smoking variable on birth weight adjusted for partner smoking, mothers age, mothers height, mothers pre-pregnancy weight, parity, and offspring sex. Dark green are the results from the individual level data MR analysis using the two-stage least squares method using G or G_2_ as the instrument, S as the exposure and birth weight is the outcome. G has no association with S = smoking before pregnancy, hence we did not perform an MR analysis. The results for applying method 1 and method 2 to estimate β_1_ − β_0_, β_1_ and β_0_ is shown in red and blue respectively.

#### 5.2.2 Exposure *S*= Smoking in the first three month of pregnancy

Mothers were asked at 16-18 weeks of gestation whether they smoked in the first three months of pregnancy. We coded those that reported ‘yes’ as *S* = 1 and mothers who reported ‘no’ as *S* = 0 and proceeded to estimate the causal effect of this exposure at the two levels (no and at least one risk allele) of rs1051730 and the genetically moderated exposure effect. In this analysis, no information about smoking before pregnancy was taken into account and therefore the mothers reporting to be non-smokers either gave up smoking when getting pregnant or did not smoke before pregnancy. Figure 6 (ii) displays the assumed DAG for our analysis. Method 1 and 2 were applied to derive estimates for *β*_1_, *β*_0_ and *β*_1_ − *β*_0_ in Table 1 and Figure 7. For these analyses, the logistic model for *S* given *G* and *G*_2_ was adjusted for whether the partner of the mother smokes, the mothers age, parity and the first 10 genetic principal components. The model predicting birth weight was adjusted for offspring sex, mothers age, mothers height, parity, mothers pre-pregnancy weight and the first 10 genetic principal components. We viewed these variables as confounders for either smoking before pregnancy or birth weight or both. For this analysis, a stronger association was observed between rs1051730 and *S*, meaning that we were cautious with the interpretation of method 2 results, given its crucial role in the in estimation of the genetically moderated exposure effect. The causal effect of smoking during pregnancy on birth weight in those with a rs1051730 risk allele, *β*_1_, was estimated to be -212g and -222g using method 1 and 2 respectively, whereas the effect of smoking in the individuals without the genetic variant on birth weight is -220g and -205g for method 1 and 2 respectively.

For both smoking exposures, we were unable to identify a difference between the two genetic groups. This could be because the effect of pre-pregnancy smoking and smoking in the first three months does not truly differ in people with or without variant rs1051730. However, a simulation investigation showed that large numbers of individuals would be required to identify a genetically driven effect of smoking with the magnitudes we observed in our analysis. Specifically, we simulated data with sample sizes from 7,000 to 500,000 and a true difference between the genetic groups of *β*_1_ − *β*_0_ between -20g and -5g. To reach 80% power in estimating *β*_1_ − *β*_0_ = −15 a sample size of 500,000 individuals is required in our simulation when using method 1. The RGMEE (method 2), which is more robust to pleiotropy compared to method 1, is able to estimate *β*_1_ − *β*_0_ = − 15 with a power of over 80% with 200,000 individuals. More details on the results of this are shown in the *Supplementary Material*. This provides important guidance on the much larger sample size, way beyond the 7752 mothers in the ALSPAC study, that would be required to detect a difference between the genetic groups in the region of what we observe here.

Despite not being able to detect a meaningful genetically moderated exposure effect, overall our results suggest that smoking before pregnancy or smoking in the first three months of pregnancy results in a lower birth weight compared to not smoking. This is in line with previous publications [26, 19, 8].

### 5.3 Observational analysis and ‘standard’ MR

In addition to applying the new proposed methods to the ALSPAC data set, we also looked at the observational association between smoking and birth weight. A linear regression of *S* (using both smoking definitions) on birth weight adjusted for partner smoking, mothers age, mothers height, mothers pre-pregnancy weight, parity and offspring sex yielded negative associational estimates. Although the observational analysis likely suffers from residual confounding, and cannot be interpreted as a causal effect, the direction of effect remains the same compared to estimating *β*_1_ and *β*_0_ (Figure 7), albeit of a smaller magnitude. As indicated in Table 1, rs1051730 (*G*) is not associated with smoking before pregnancy. Therefore, we are unable to perform a standard MR analysis using *G* as the genetic instrument for *S* = smoking before pregnancy. However, we did perform a standard MR analysis using individual level data and the two-stage least squares approach with *S* = smoking in the first three months of pregnancy. We explain in Section 2.1 that the standard MR with a homogeneity violating instrument like rs1051730 estimates the CACE (while the monotonicity assumption holds). The CACE of smoking in the first the months of pregnancy on birth weight is -210 grams (95% CI:(−293,-128)). Note that these result potentially suffer from weak instrument bias.

Additionally, and as a confirmatory test of our previous genetic analyses we calculated a standard MR estimate using the genetic risk score for smoking initiation to instrument smoking before and smoking in the first three months of pregnancy. For these analyses rs1051730 is not considered. The methodology is described in Section 3.3. Using equation 6 the estimated *β*-values obtained from applying method 1 and 2 to the ALSPAC data we derive an average causal effect of -165g for smoking before pregnancy. This compares to the average causal effect of -164g (Figure 7) estimated with standard MR approach. Similarly, for smoking in the first three months of pregnancy we obtain an estimate of -213 g using the *β*-values from method 1 or method 2 outputs and the formula provided above.

## 6 Discussion

In this paper we propose a general framework for MR that allows the inclusion of traditional genetic instruments, as well as those that violate the key homogeneity assumption. This enables an analysis that goes beyond estimation of the average causal effect to consider estimation within specific genetic sub-groups, with a view to quantifying genetically driven effect heterogeneity. Our approach builds on ideas from the pharmacogenetic TWIST framework proposed by Bowden et al. [5] to a more mainstream epidemiological setting, as well as incorporating the technique of multivariable MR [27]. Specifically, method 1 offers a new approach to estimate the genetically moderated exposure effect, which could be triangulated with existing methods. Furthermore, method 1 allows for a direct effect between the homogeneity-violating instrument and the exposure. In the presence of unmeasured confounding, the methods proposed in the paper by Bowden et al. [5] require the instrument to be independent of the exposure. To allow for a direct pleiotropic effect between the homogeneity-violating instrument and the outcome we proposed method 2. Simulation studies revealed the necessary sample sizes to detect an effect with sufficient power under method 1 and 2, considering plausible genetically moderated exposure effect sizes. To motivate the methods, we applied them to data from the ALSPAC cohort to investigate the effect of smoking before and in pregnancy on birth weight using a traditional genetic risk score for smoking and a rs1051730 as an effect modifier.

Our work could be further extended by considering the incorporation of additional methods to allow for the relaxation of further key assumptions. For example, allowing a genetic variant with a pleiotropic effect that acts through an unmeasured confounder (i.e. correlated pleiotropy). We assumed that the underlying data structure follows a linear interaction model. Future work could explore different data structures and non-linear effects. For simplicity, and to naturally follow on from the approach proposed in Bowden et al. we assumed a binary effect modifying instrument through the dichotomization of the genetic instrument rather than using the number of risk alleles. Future work could relax this assumption.

Despite not being able to show a difference between the two genetic groups in our applied example due to a limitation in sample size, our investigation clarifies how large future cohort study samples need to be to estimate effects of the magnitude we observed. We believe our framework is a useful methodological extension to investigate genetically driven heterogeneity. Our methods could for example be applied in other setting where larger sample sizes are available, or by meta-analysing results with additional cohorts. Settings outside of pregnancy research are also possible and would not require mother and child pair information. For example, investigating the genetically driven effect of continuing smoking on lung cancer. Other examples could be using genetic variants associated with reducing alcohol consumption and the effects on various health outcomes. Data sets like the UK Biobank with genetic information available for 500,000 individuals could be used. Software for implementing the methods can be found at https://github.com/AJaitner/paper_heterogeneity_MR[to-be-added] as well as code to implement the simulation studies and applied analysis.

## 7 Declarations

### 7.1 Ethics approval

Ethical approval for the study was obtained from the ALSPAC Ethics and Law Committee and the Local Research Ethics Committees. Informed consent for the use of data collected via questionnaires and clinics was obtained from participants following the recommendations of the ALSPAC Ethics and Law Committee at the time.

### 7.2 Data Availability

The data in ALSPAC is fully available, via managed systems, to any researchers. The managed system is a requirement of the study funders, but access is not restricted on the basis of overlap with other applications to use the data or on the basis of peer review of the proposed science. The ALSPAC data management plan describes in detail the policy regarding data sharing, which is through a system of managed open access. The following steps highlight how to apply for access to the data included in this paper and all other ALSPAC data. (1) Please read the ALSPAC access policy, which describes the process of accessing the data and samples in detail and outlines the costs associated with doing so. (2) You may also find it useful to browse the fully searchable ALSPAC research proposals database, which lists all research projects that have been approved since April 2011. (3) Please submit your research proposal for consideration by the ALSPAC Executive Committee. You will receive a response within 10 working days to advise you whether your proposal has been approved. If you have any questions about accessing data, please.

## 7.5 Acknowledgements

We are extremely grateful to all the families who took part in this study, the midwives for their help in recruiting them, and the whole ALSPAC team, which includes interviewers, computer and laboratory technicians, clerical workers, research scientists, volunteers, managers, receptionists and nurses.

The authors would like to acknowledge the use of the University of Exeter High-Performance Computing (HPC) facility in carrying out this work.

## 8 Appendix: MR estimate equals the Complier Average Causal Effect if Homogeneity is violated but Monotonicity holds

To better understand the bias term of (5) from Section 2.1 in the main paper in the presence of IV4 violation, we first introduce the Principle Stratum framework described in Angrist, Imbens, and Rubin [28]. In our context, we imagine the existence of four compliance classes:

- *Compliers*: Individuals that smoke if they have the risk allele (*G* = 1) and do not smoke if they do not have it (*G* = 0).
- *Always Smokers*: Individuals that always smoke regardless of their genotype.
- *Never Smokers*: Individuals that never smoke regardless of their genotype.
- *Defiers*: Individuals that go against their genotype, this means they smoke if they do not have the risk allele and do not smoke if they do have the risk allele.

Formally this can be written as in Table 2 which relates compliance classes (and their proportion in the population) to the joint values of two potential smoking variables, *S*(*G* = 1) and *S*(*G* = 0) [29]:

**Table 2:** Relating compliance classes (and their proportion in the population) to the joint values of two potential smoking variables.

| | $S(G = 1)$ | $S(G = 0)$ | Proportion |
| --- | --- | --- | --- |
| Compliers (c) | 1 | 0 | $\pi_c$ |
| Always Smokers (as) | 1 | 1 | $\pi_{as}$ |
| Never Smokers (ns) | 0 | 0 | $\pi_{ns}$ |
| Defiers (d) | 0 | 1 | $\pi_d$ |

If we assume IV4 is violated but an alternative assumption, that there are no *Defiers* (*π*_*d*_ = 0, also termed ‘Monotonicity’), then from Table 2, we can equate:

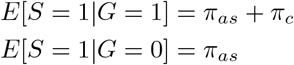

Furthermore, *β*_1_ in equation (5) reflects the average causal effect of smoking amongst those who smoke, which is the union of the compliers and always smokers and *β*_0_ reflects the effect of smoking in the always smokers only (due to the assumptions of no defiers). We can express *β*_1_ itself as a weighted average of the causal effect of smoking in compliers (*β*_*c*_) and always smokers (*β*_0_):

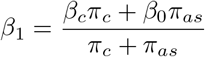

Through re-arrangement of the above expression, equation (5) can then be written as:

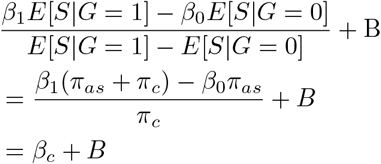

Therefore, if Homogeneity is violated, but Monotonicity holds, MR targets the average causal effect of smoking among *Compliers* (the complier average causal effect(CACE)) plus any bias due to violation of IV2 and IV3.

## 9 Appendix: Mean and standard derivation across simulated and ALSPAC data sets

In Table 3 the mean and standard derivation for each variable are shown for the ALSPAC data set (n = 7752) and the different simulation data sets. Depending on the definition of smoking the results are slightly different. Therefore, we show the mean and variance for ALSPAC defining *S* as smoking before pregnancy (yes/no) and for smoking in the first three month of pregnancy (yes/no). As described in the main body of the paper, we simulate data sets with different sample sizes and applied all methods to them. For each method we assume that the required assumptions for the respective method hold. This means that the data generating model is slightly different for each method and this also reflects in the mean and standard derivation for each variable. As an example we show the mean and variance for each variables for the different methods for a data set with 50,000 individuals. Note that each simulation is repeated 20,000 times and therefore for each method 20,000 data sets are generated. Hence, the mean value of the 20,000 mean values is displayed (similarly for the standard derivation).

**Table 3:** Mean and standard derivation of the outcome, smoking variable and rs1051730 in ALSPAC (n = 7752) and in simulated data sets with 50,000 individuals.

| Data set | $\bar{Y} (\sigma(Y))$ | $\bar{S} (\sigma(S))$ | $\bar{G} (\sigma(G))$ |
| --- | --- | --- | --- |
| ALSPAC, S = pre-pergnnacy smoking | 3474.21 (476.87) | 0.31 (0.46) | 0.55 (0.50) |
| ALSPAC, S = smoking in the 1st 3 month | 3474.21 (476.8741) | 0.23 (0.42) | 0.55 (0.50) |
| Method 1 simulation $n= 50,000$ | 3459.17 (476.86) | 0.26 (0.44) | 0.55 (0.50) |
| Method 2 simulation $n= 50,000$ | 3506.07 (477.35) | 0.24 (0.43) | 0.55 (0.50) |

## 10 Appendix: Simulation parameter values for each method

In order for the assumptions for each method to be satisfied, the data generating model needs to be slightly different. Here we show the parameter values used for the data generation for each method.

**Table 4:** Simulation parameter values to generate simulated data for method 1 and method 2.

| Method | $\gamma_{UG}$ | $\gamma_{SG}$ | $\gamma_{SG_2}$ | $\gamma_{SU}$ | $\gamma_{YG}$ | $\gamma_{YU}$ | $\beta_1$ | $\beta_0$ |
| --- | --- | --- | --- | --- | --- | --- | --- | --- |
| 1 | 0 | 0.2 | 0.8 | 1.6 | 0 | 80 | -200 | -100 |
| 2 | 0 | 0 | 0.8 | 1.6 | 80 | 80 | -200 | -100 |

## 11 Appendix: Simulations to show which assumptions need to hold for each method

We simulated data following the data generation process described in section 4.1. We aimed to match the parameters of the data generating model as close to the ALSPAC data set as possible. We choose to set *β*_1_ = −200 and *β*_0_ = −100, which assumes a genetically moderated effect of *β*_1_ − *β*_0_ = −100 and a twice as large effect in the *G* = 1 group compared to the *G* = 0 group. We chose scenarios where some of the assumptions hold and others do not hold. For all simulations we choose a sample size of *n* = 20, 000 and repeated each simulation 20,000 times.

### 11.1 Simulation for method 1

We investigated 9 different scenarios and show the density plots for the estimation of *β*_1_ and *β*_0_. We expect unbiased estimations for scenario 1-6 and verify that method 1 is robust to whether there is a direct effect between *G* and *S*. Scenario 7-8 are biased due to direct and/or indirect pleiotropy of the genetic instrument *G*. The density plots over the 20,000 simulation runs for each scenario are shown in Figure 8. As expected, strength of the instrument influences the precision of the estimation and therefore we see in Figure 8 that the density peak is higher for scenario 3 and scenario 6 compared to the other scenarios. It is also clearly visible that the scenarios with pleiotropy of the gentic instrument *G* (scenario 7-9) lead to biased results.

**Figure 8:**
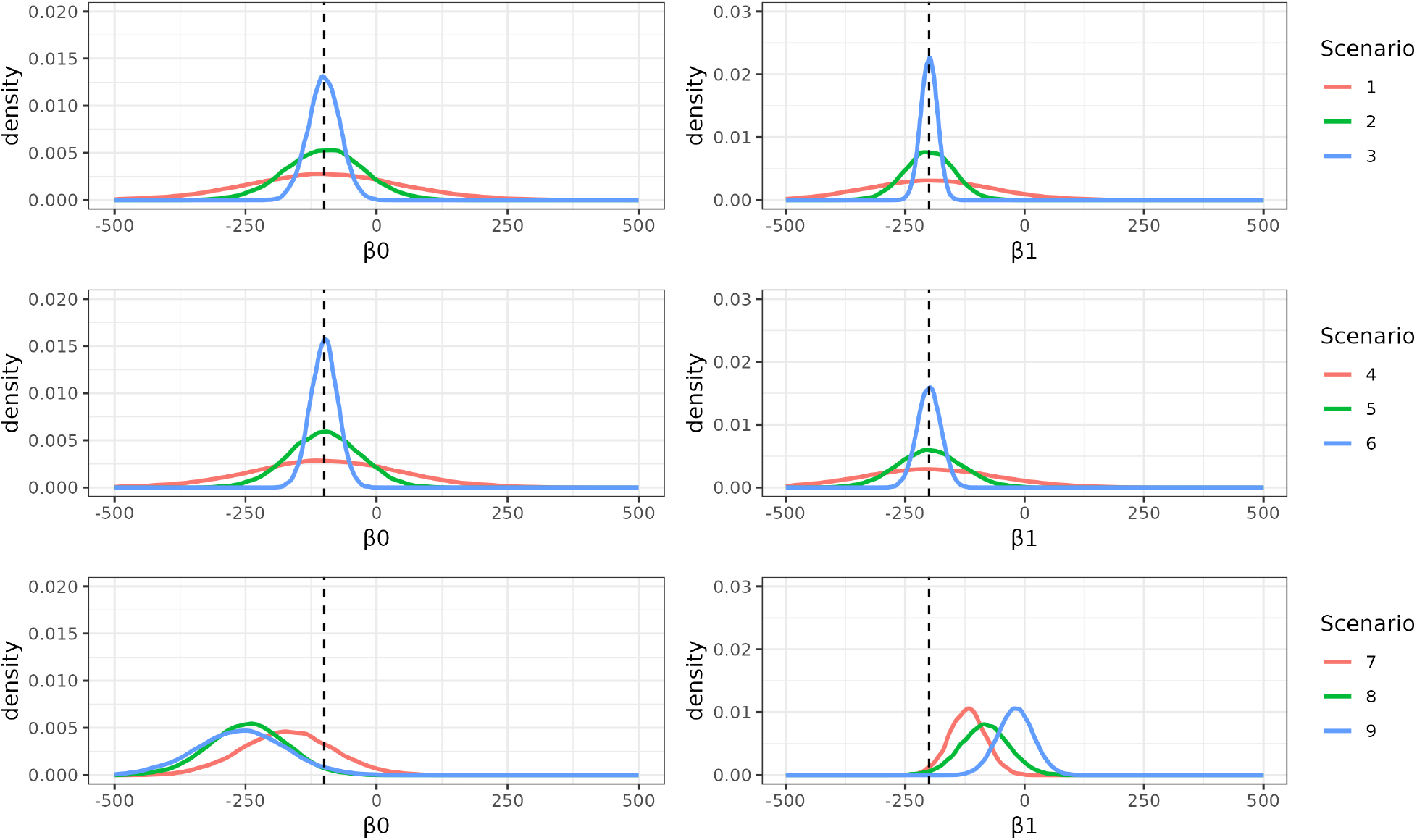
Density plots for the estimation of β_0_ and β_1_ using method 1 over 20,000 simulation runs for 9 different scenarios.

**Table 5:** Parameter values for the difference scenarios to verify the assumptions for method 1. Increasing strength of the instrument G and G_2_ for scenario 1-3 and increasing strength of the instrument for G for scenario 4-6. The confounder parameters are set to γ_Y U_ = 80, γ_SU_ = 1.6.

| | Description | $\gamma_{SG}$ | $\gamma_{SG_2}$ | $\gamma_{YG}$ | $\gamma_{UG}$ | F-Stat $G$ | F-Stat $G_2$ |
| --- | --- | --- | --- | --- | --- | --- | --- |
| 1 | Direct effect between $G$ and $S$ | 0.2 | 0.4 | 0 | 0 | 71 | 19 |
| 2 | Direct effect between $G$ and $S$ | 0.8 | 0.8 | 0 | 0 | 309 | 302 |
| 3 | Direct effect between $G$ and $S$ | 2 | 2 | 0 | 0 | 1765 | 1818 |
| 4 | No direct effect between $G$ and $S$ | 0 | 0.4 | 0 | 0 | 69 | 1 |
| 5 | No direct effect between $G$ and $S$ | 0 | 0.8 | 0 | 0 | 286 | 1 |
| 6 | No direct effect between $G$ and $S$ | 0 | 2 | 0 | 0 | 1850 | 1 |
| 7 | Indirect pleiotropy | 0.8 | 0.8 | 0 | 0.8 | 310 | 2013 |
| 8 | Direct pleiotropy | 0.8 | 0.8 | 80 | 0 | 309 | 302 |
| 9 | Indirect and direct pleiotropy | 0.8 | 0.8 | 80 | 0.8 | 310 | 2014 |

### 11.2 Simulation for method 2

We investigated 8 different scenarios and show the density plots for the estimation of *β*_0_ and *β*_1_ − *β*_0_. We expect only scenario 1 and 2 to be unbiased. The direct effect between *S* and *G* bias the estimation of *β*_1_ −*β*_0_ and as this estimation is crucial for the following steps of estimating *β*_0_. However, as the biased estimation is not a visible in Figure 9 we also provide the mean and standard derivation for the estimates for each scenario in Table 7. An effect between the genetic instrument *G* and the unmeasured confounder *U* and hence the indirect pleiotropy results in biased estimates for scenario 3-8. However, a direct pleiotropic effect of *G* is no problem (scenario 2 is unbiased)

**Table 6:** Parameter values for the difference scenarios to verify the assumptions for method 2. The confounder parameters are set to γ_Y U_ = 80, γ_SU_ = 1.6.

| | Description | $\gamma_{SG}$ | $\gamma_{SG_2}$ | $\gamma_{YG}$ | $\gamma_{UG}$ |
| --- | --- | --- | --- | --- | --- |
| 1 | No direct effect between $G$ and $S$, no pleiotropy | 0 | 0.8 | 0 | 0 |
| 2 | No direct effect between $G$ and $S$, direct pleiotropy | 0 | 0.8 | 80 | 0 |
| 3 | No direct effect between $G$ and $S$, indirect pleiotropy | 0 | 0.8 | 0 | 0.8 |
| 4 | No direct effect between $G$ and $S$, indirect and direct pleiotropy | 0 | 0.8 | 80 | 0.8 |
| 5 | Direct effect between $G$ and $S$, no pleiotropy | 0.8 | 0.8 | 0 | 0 |
| 6 | Direct effect between $G$ and $S$, direct pleiotropy | 0.8 | 0.8 | 80 | 0 |
| 7 | Direct effect between $G$ and $S$, indirect pleiotropy | 0.8 | 0.8 | 0 | 0.8 |
| 8 | Direct effect between $G$ and $S$, indirect and direct pleiotropy | 0.8 | 0.8 | 80 | 0.8 |

**Table 7:**
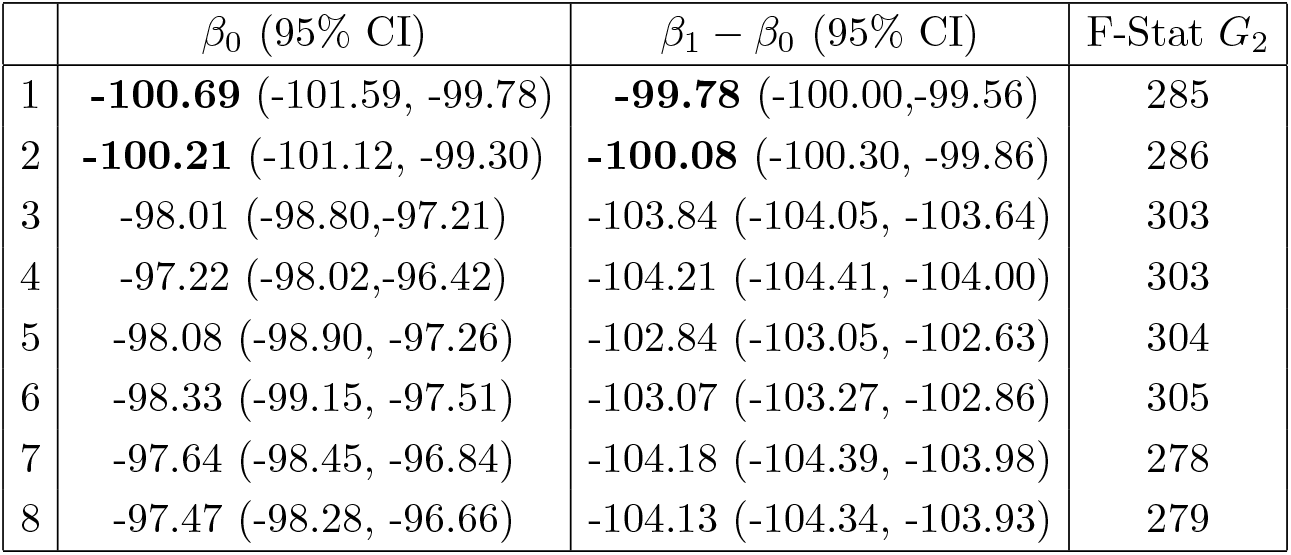
Mean values and 95% confidence intervals for the estimation of β_0_ and β_1_ − β_0_ using method 2 for different scenarios. Estimates for scenario 1 and 2 are highlighted in bold because the true value (−100) lies within the 95 % confidence interval.

| | $\beta_0$ (95% CI) | $\beta_1 - \beta_0$ (95% CI) | F-Stat $G_2$ |
| --- | --- | --- | --- |
| 1 | **-100.69** (-101.59, -99.78) | **-99.78** (-100.00, -99.56) | 285 |
| 2 | **-100.21** (-101.12, -99.30) | **-100.08** (-100.30, -99.86) | 286 |
| 3 | -98.01 (-98.80, -97.21) | -103.84 (-104.05, -103.64) | 303 |
| 4 | -97.22 (-98.02, -96.42) | -104.21 (-104.41, -104.00) | 303 |
| 5 | -98.08 (-98.90, -97.26) | -102.84 (-103.05, -102.63) | 304 |
| 6 | -98.33 (-99.15, -97.51) | -103.07 (-103.27, -102.86) | 305 |
| 7 | -97.64 (-98.45, -96.84) | -104.18 (-104.39, -103.98) | 278 |
| 8 | -97.47 (-98.28, -96.66) | -104.13 (-104.34, -103.93) | 279 |

**Figure 9:**
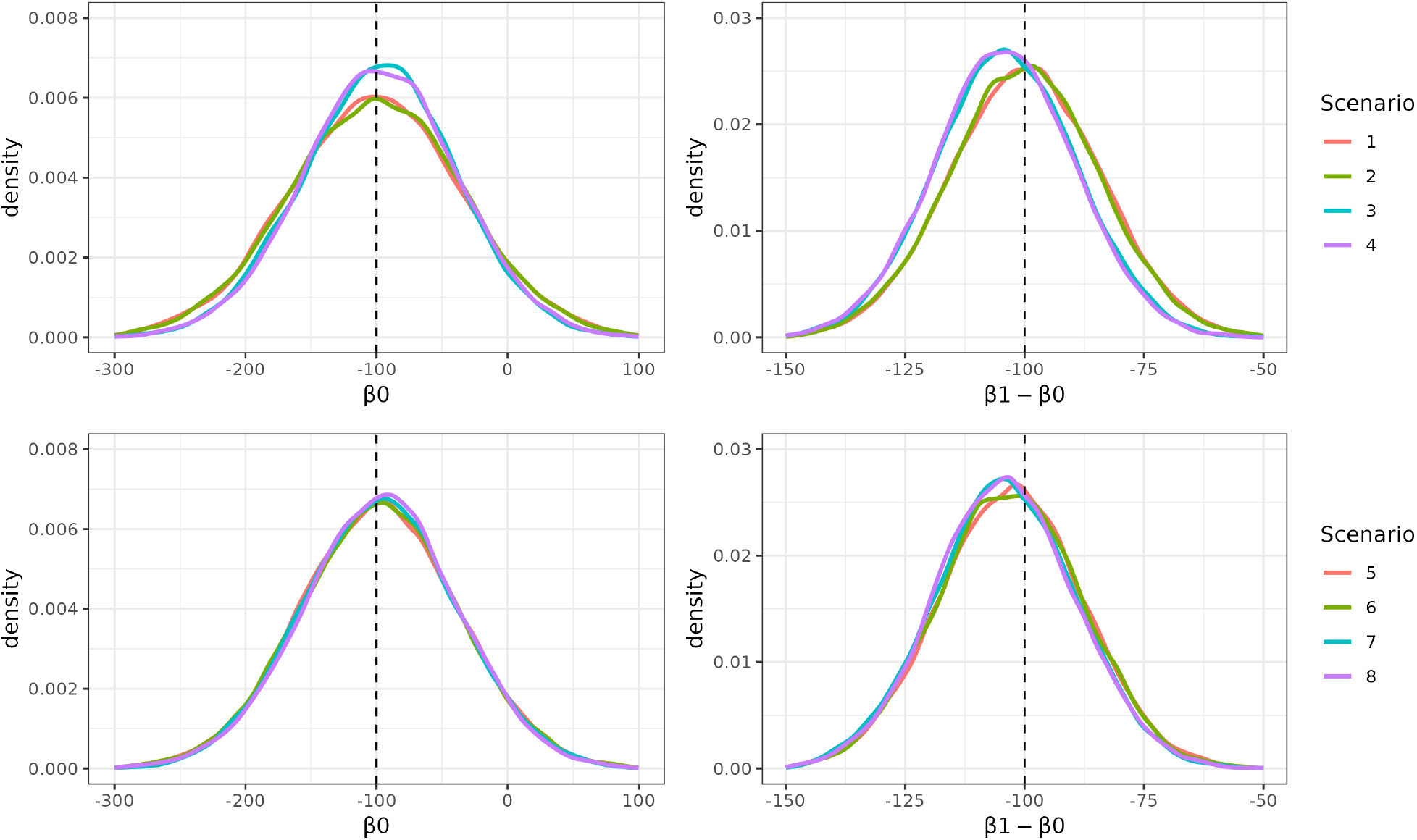
Density plots for the estimation of β_0_ and β_1_ − β_0_ using method 2 over 20,000 simulation runs for 8 different scenarios.

## 12 Appendix: Simulation of power for small differences between genetic groups

To investigate at which sample size the two methods are able to estimate a small *β*_1_ − *β*_0_ with sufficient power, we simulated different data sets. The data generation follows the same procedure as described in the main body of this paper. The parameter values *γ* are kept the same as in previous simulations. We investigated 4 different scenarios as shown in Table 8. Each scenario was simulated with sample sizes varying from 7,000 to 500,000. Each simulation was repeated 5,000 times (*N* = 5, 000). We estimate the power to reject the null hypothesis that *β*_1_ − *β*_0_ were statistically different from zero at the 5% significance level across the 5,000 simulations. The results for both methods are shown in Figure 10.

**Table 8:** Four different scenarios with small values for β_1_ − β_0_, for data simulations with different large sample sizes.

| Scenario | $\beta_1$ | $\beta_0$ | $\beta_1 - \beta_0$ |
| --- | --- | --- | --- |
| 1 | -220 | -200 | -20 |
| 2 | -200 | -185 | -15 |
| 3 | -180 | -170 | -10 |
| 4 | -168 | -163 | -5 |

**Figure 10:**
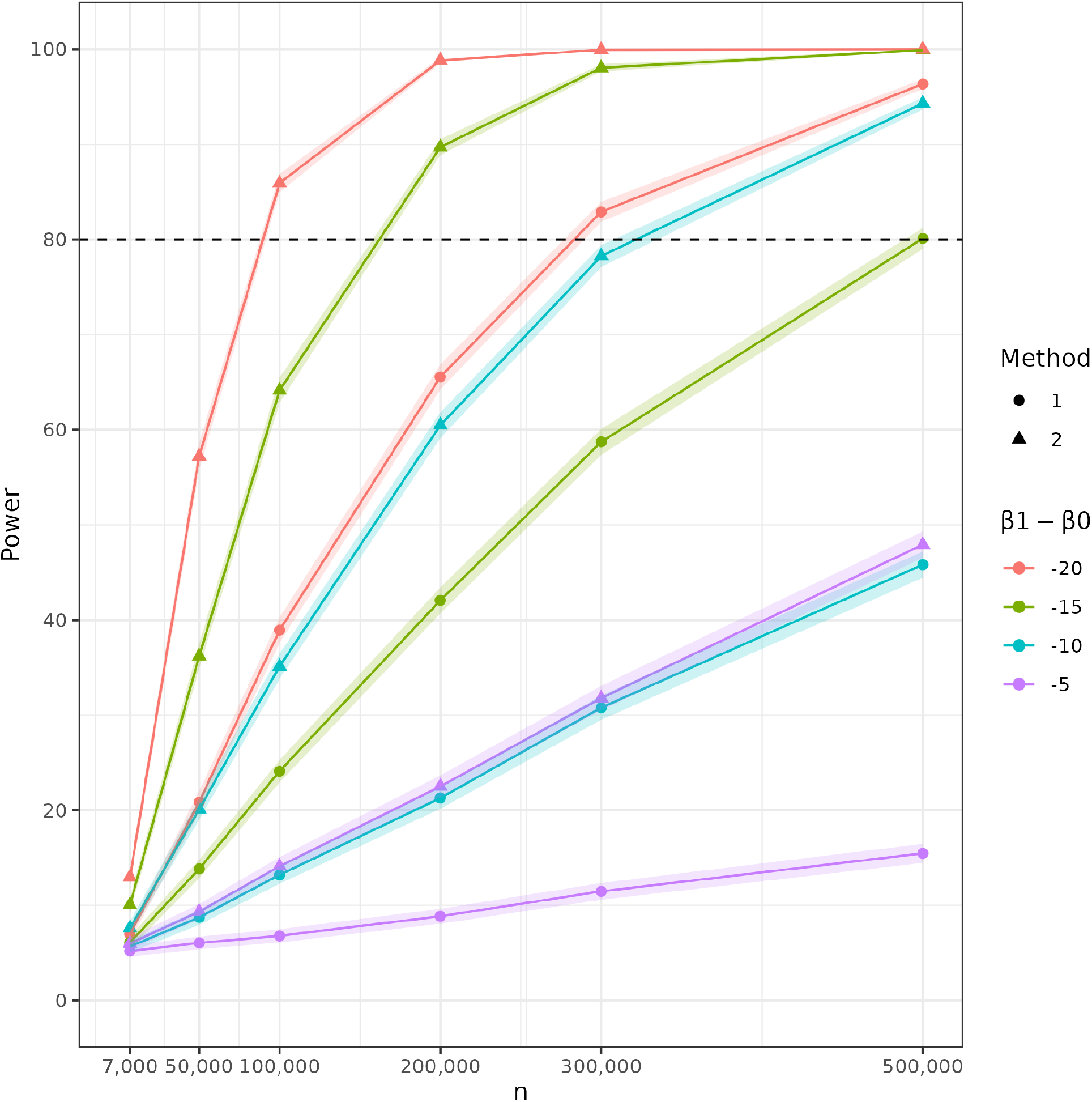
Power to estimate β_1_ −β_0_ for different samples sizes (on the x-axis) applying method 1 and method 2 (see different shapes). The different colours refer to the different simulated data sets with changing β_1_ − β_0_ values.

